# A Preliminary Investigation Of Repetitive Transcranial Magnetic Stimulation Applied To The Left Dorsolateral Prefrontal Cortex In Treatment Seeking Participants With Cannabis Use Disorder

**DOI:** 10.1101/2023.07.10.23292461

**Authors:** Gregory L. Sahlem, Bohye Kim, Nathaniel L. Baker, Brendan L. Wong, Margaret A. Caruso, Lauren A. Campbell, Irakli Kaloani, Brian J. Sherman, Tiffany J. Ford, Ahmad H. Musleh, Jane P. Kim, Nolan R. Williams, Andrew J. Manett, Ian H. Kratter, Edward B. Short, Terese K. Killeen, Mark S. George, Aimee L. McRae-Clark

**Affiliations:** Department of Psychiatry and Behavioral Sciences, Stanford University, Palo Alto, California, USA Departments of; Public Health Sciences; Psychiatry, Medical University of South Carolina, Charleston, South Carolina, USA; Ralph H. Johnson Veterans Administration Medical Center, Charleston, South Carolina, USA

**Keywords:** Cannabis use disorder, marijuana, cannabis, addiction, TMS, transcranial magnetic stimulation

## Abstract

**Background:** Cannabis use disorder (CUD) is a common and consequential disorder. When applied to the dorsolateral prefrontal cortex (DLPFC), repetitive transcranial magnetic stimulation (rTMS) reduces craving across substance use disorders and may have a therapeutic clinical effect when applied in serial sessions. The present study sought to preliminarily determine whether serial sessions of rTMS applied to the DLPFC had a therapeutic effect in CUD.

**Methods:** This study was a two-site, phase-2, double-blind, randomized-controlled-trial. Seventy-two treatment-seeking participants (37.5% Women, mean age 30.2±9.9SD) with ≥moderate-CUD were randomized to active or sham rTMS (Beam-F3, 10Hz, 20-total-sessions, with cannabis cues) while undergoing a three-session motivational enhancement therapy intervention. The primary outcome was the change in craving between pre- and post- treatment (Marijuana Craving Questionnaire Short-Form—MCQ-SF). Secondary outcomes included the number of weeks of abstinence and the number of days-per-week of cannabis use during 4-weeks of follow-up.

**Results:** There were no significant differences in craving between conditions. Participants who received active rTMS reported numerically, but not significantly, more weeks of abstinence in the follow-up period than those who received sham rTMS (15.5%-Active; 9.3%-Sham; rate ratio = 1.66 [95% CI: 0.84, 3.28]; *p*=0.14). Participants who received active rTMS reported fewer days-per-week of cannabis use over the final two-weeks of the follow-up period (Active vs. Sham: -0.72; Z=-2.33, *p*=0.02).

**Conclusions:** This trial suggests rTMS is safe and feasible in individuals with CUD and may have a therapeutic effect on frequency of cannabis use, though further study is needed with additional rTMS-sessions and a longer follow-up period.

**Highlights:** This phase-2 RCT tested the efficacy of prefrontal rTMS for cannabis use disorder

The study paradigm was safe and feasible, and participants tolerated rTMS well

The active-group had numerically more weeks of abstinence during follow-up

The active-group had fewer days-per-week of cannabis use during follow-up

More rTMS and a longer follow-up may result in a larger effect in future studies

## 1.0 Introduction

Cannabis Use Disorder (CUD) is a common condition with well-documented adverse effects^1–3^ and concordantly high demands for treatment^4,5^. The incidence of frequent cannabis use and CUD may be increasing in the United States and worldwide^6,7^ with increasing legalization and decreased perceived risk^8^. The frequency of daily cannabis use has also risen in recent years in the United States—potentially further increasing the risk of an increased incidence of CUD in the future^9^. Though there are promising pharmacologic treatments in the pipeline^10–13^, no medication has distinguished itself as clearly effective in the treatment of CUD. Further, although consistently demonstrating a beneficial effect, studies testing behavioral therapies for CUD have resulted in moderate effects^14^. As such, there remains a need to develop new therapeutics for CUD.

Repetitive transcranial magnetic stimulation (rTMS) works via the principles of magnetic induction and long-term potentiation^15–17^ and can focally alter circuit function in the brain^18–20^. Trials applying serial applications of rTMS in a variety of neuropsychiatric conditions have demonstrated that by varying the location of stimulation and the treatment paradigm, it is possible to derive a therapeutic benefit in different illnesses, and rTMS is now cleared by the US Food and Drug Administration for the treatment of Major Depressive Disorder^21–23^, Obsessive Compulsive Disorder^24^, and Tobacco Use Disorder^25^. In line with the several indications for treatment, there has been increasing promise that rTMS may become a therapeutic option across addictions, including CUD^26^. Studies have suggested rTMS has the potential to effect behavioral aspects of addiction^27–30^, engage its neurocircuitry^20,31,32^, and, when serial sessions of rTMS are applied, have therapeutic effects. Several neurocircuit targets have emerged for study in therapeutic trials, with early promising results for the left dorsolateral prefrontal cortex (DLPFC)^33^, the frontal pole^34^, the dorsomedial prefrontal cortex^35^, and the anterior-insula / inferior frontal gyrus^25^, although not all trials have resulted in a beneficial effect^36^.

Our group and others^37^ have explored the potential effect of applying rTMS to the left DLPFC, first in non-treatment-seeking participants with CUD^38^ and then participants with CUD who were interested in reducing their use of cannabis^39^. Our early findings suggested that a single-session of rTMS could be feasibly applied to participants with CUD, was generally well tolerated, and may reduce the purposefulness aspect of craving^38^. In a subsequent study, we found that it was infeasible to deliver daily sessions of rTMS for two-weeks in treatment-seeking participants with CUD. However, those participants who did attend daily sessions reported less craving and reduced cannabis use that persisted 4-weeks after receiving rTMS^39^. The findings from our preliminary work and other therapeutic studies in other addictive disorders applying rTMS to the DLPFC, suggested therapeutic promise for CUD, albeit with a treatment paradigm that differed from daily applications. Both data^40–42^ and clinical experience suggest it is possible to get a therapeutic effect using rTMS even if treatments are delivered less frequently than daily. Based on both trial experience and qualitative discussions with participants from our pilot treatment trial, we hypothesized that delivering study-treatments twice each week would be feasible and have a clinical effect. We subsequently designed the present phase-2 study to preliminarily determine if rTMS applied to the DLPFC twice-weekly had the potential to help treatment-seeking participants with CUD reduce their cannabis use. Specifically, we hypothesized that participants receiving active-rTMS would have reduced craving and more weeks of abstinence than participants receiving sham-rTMS.

## 2.0 Materials and Methods

### 2.1 Study Design

This study was an outpatient, two-site, double-blind, randomized, parallel-designed, sham-controlled trial. Two sites conducted the trial (initially at the Medical University of South Carolina between August 2017 and March 2020, and, subsequently, at Stanford University between April 2021 and June 2022). There was a 1:1 allocation to active or sham rTMS without stratifying variables. Participants were evaluated at a screening visit and underwent study-treatment over five-weeks, where they attended two study-treatment-visits per week (and received two study-rTMS-treatments per visit) for twenty total study-treatments delivered over ten study-treatment-visits. Participants met with a study therapist during three of the study visits and received Motivational Enhancement Therapy (MET)^13,43^. Participants returned two- and four-weeks after the final study-treatment-visit to complete follow-up assessments. We compensated participants for their time and travel and used prize-based contingency management to reinforce visit attendance. This trial was conducted in accordance with the Declaration of Helsinki, was approved by the Institutional Review Boards of each study site, and was pre-registered on clinicaltrials.gov (NCT03144232). Participants provided written informed consent before engaging in study-procedures.

### 2.2 Participant Selection

We recruited participants from addiction medicine clinics and from the community via media advertisements. Participants were included if: a) they were between the ages of 18 and 60 years; b) they met the Diagnostic and Statistical Manual for Mental Disorders—DSM-5–criteria for ≥moderate cannabis use disorder; c) they had a desire to quit or reduce cannabis use; and d) they had a positive urine drug test for cannabis. Participants were excluded if: a) they were pregnant or breast-feeding; b) they met DSM-5-criteria for another ≥moderate substance use disorder (other than nicotine use disorder); c) they were regularly taking medications with central nervous system effects; d) they had a history of psychotic disorder, bipolar disorder, or any other psychiatric condition requiring acute treatment; e) they had a Hamilton Rating Scale for Depression—HRSD_24_ score greater than 10 indicating clinically relevant depressive symptoms; f) they had a history of dementia or other cognitive impairment; g) they had active suicidal ideation or a suicide attempt within the past 90-days; h) they had any contraindications to receiving rTMS^44,45^ or MRI^46^, or; i) they had any unstable general medical condition.

### 2.3 Assessments

Participants were allowed to use cannabis ad libitum prior to the screening visit, though they were instructed not to arrive intoxicated (verified via clinical assessment). During the screening visit, participants underwent a medical and psychiatric evaluation which included the Mini-International Neuropsychiatric Interview (MINI)^47^, the Hamilton Rating Scale for Depression (HRSD_24_)^48^, and the structured criteria for CUD as found in the Structured Clinical Interview for the DSM-5^49^. We quantified CUD severity using the Cannabis Use Disorder Identification Test (CUDIT)^50^ and assessed motivation to change using the Marijuana Contemplation Ladder^51^. We collected the Marijuana Problem Scale as part of the Brief Marijuana Dependence Counseling Personal Feedback Report^43^. We assessed current symptoms of cannabis withdrawal using the Cannabis Withdrawal Scale (CWS)^52^. We recorded twenty-eight days of previous cannabis use using the Time-Line Follow-Back (TLFB)^53^. We defined a cannabis use session as any cannabis use separated by an hour or more since the last cannabis use session and approximated the number of grams used at each session. We also assessed alcohol and other substance use using qualitative urine drug testing including testing for ethyl-glucuronide (T-Cup®, Guangzhou Wondfo Biotech Co., LTD).

Eligible participants received rTMS during ten study-visits. Participants were instructed to abstain from cannabis and other substances for at least 24-hours prior to rTMS visits #1 and #10 (verified by saliva drug testing; SalivaConfirm® testing, Confirm Biosciences, Inc.) and we collected immediate pre- and post- data on those visits. Participants completed a visual cue-reactivity task^54^ during functional magnetic resonance imaging (fMRI) at the beginning of the pre- and post- visits (results will be reported elsewhere) and then we measured craving using the Short-Form of the Marijuana Craving Questionnaire (MCQ-SF)^55^ approximately 20-minutes later. Additionally, data was collected for cannabis use via the TLFB (preceding week) and symptoms of withdrawal via the CWS (after 24-hours of abstinence). We used the Architect C4000 system from Abbott Laboratories to measure urine cannabinoids and creatinine, and the creatinine-corrected urine cannabinoid level (ng/mg) was derived by dividing the cannabinoid level (ng/ml) by the creatinine level (mg/ml).

The remainder of the study-visits were conducted while participants were allowed to use cannabis ad libitum (though were not intoxicated during visits). We collected TLFB data continuously and assessed symptoms of withdrawal using the CWS at each visit. Participants met with the study team two- and four-weeks following their last rTMS-visit, during which cannabis use was assessed for the 4-weeks of follow-up using the TLFB and urine cannabinoids.

We defined weeks of abstinence from cannabis as those weeks where participants did not report any cannabis use. On the visits where urine cannabinoids were collected, self-reported weeks of abstinence were verified as abstinent, defined in^56^ as a 25% drop in creatinine-corrected cannabinoid level from the prior level and an absolute level of <200ng/ml.

We identified adverse events at each visit using open-ended questions and considered them in the context of their severity, seriousness, and possible relation to the study-intervention.

### 2.4 Repetitive Transcranial Magnetic Stimulation (rTMS) Paradigm

Participants received active or sham rTMS via a MagVenture MagPro X100 double-blinded device using a B65 coil. During each session, participants received 4000 pulses of stimulation at 10Hz (5-seconds on, 10-seconds off) while interacting with cannabis cues^57^. We delivered stimuli at 120% of the participants resting motor threshold^58^. We targeted the left dorsolateral prefrontal cortex using the Beam-F3 method^59^. We delivered two-sessions of rTMS at each of the ten study-treatment-visits with a 30-minute inter-session-interval. Sham stimulation consisted of low current electrical stimuli coinciding with each click of the TMS coil. We assessed blinding following the first and last study-session by asking participants to guess their treatment allocation and to rate their confidence on a Likert scale.

### 2.5 Study Hypotheses (Supplemental Figure-1)

The primary hypothesis (Aim-1) of the experiment, as pre-specified on clinicaltrials.gov, was that the active-rTMS-group would have a reduction in behavioral craving (via the MCQ-SF) between the immediate pre- and post- visits relative to the sham-rTMS-group. We chose immediate pre- and post- time-points (rather than longitudinal craving) because those assessments were completed with at least 24-hours of abstinence from cannabis.

The primary clinical exploratory analysis (Aim-2a), as pre-specified on clinicaltrials.gov, was that the active rTMS group would have more weeks of urine cannabinoid-verified abstinence in the follow-up period than the sham rTMS group. We selected the four-weeks of follow-up as the time frame given it followed the delivery of the entire course of rTMS and would be less subject to any supportive clinician effect that twice-weekly meetings might have. We were able to confirm abstinence using creatinine-corrected urine cannabinoids at the two- and four- week follow-up visits. We additionally tested the hypothesis that the active-rTMS-group would have fewer days-per-week of cannabis use in the follow-up period than the sham rTMS-group (Aim-2b), though this was an exploratory hypothesis that was not pre-specified. We chose abstinence and days-per-week of cannabis use as the clinical outcomes because they are not influenced by the methodologic problems inherent in assessing cannabis use (variable THC concentrations, amount used per cannabis-use-session, varying routes of administration, etc.). Further, weeks of abstinence and days-per-week of cannabis use have been associated with improved quality of life, reduced marijuana-related problems, and decreased symptoms of depression and anxiety^60–62^.

### 2.6 Analysis Plan

We calculated descriptive statistics such as means and standard deviations (SD) for all continuous variables and proportions for categorical variables. We compared demographic, screening, blinding, and adverse event variables (compiled using MEDra criteria) across condition and site using t-tests or the Wilcoxon Rank Sum test for continuous variables and the Chi-Square or Fisher’s Exact test for categorical variables.

For Aim-1, we utilized a generalized linear mixed-effects model (GLMM) to perform the analyses, with the full ITT sample (N=72) and the total MCQ-SF score as the dependent measure. We used two types of a priori models for each analysis: 1) a three-way interaction model which included an interaction between treatment, site, and time, as well as the main effects of treatment, site, time, and all pairwise interactions between the main terms, and 2) a two-way interaction model which included treatment, time, an interaction between treatment and time, and site. We considered pre- and post- variables with a strong clinical rationale for inclusion in covariate adjustment including the amount of time since the last cannabis use, CWS score, the days-per-week of cannabis use, the grams of cannabis used per day, and the number of cannabis use sessions per day.

To assess group differences in the number of weeks of abstinence in the follow-up period (Aim-2a), a Poisson regression model was fit with the total number of weeks of self-reported abstinence in the four-week follow-up period as the dependent variable and treatment, site, and treatment by site interaction as independent variables. We conservatively imputed missing values as non-abstinent, thereby including the total ITT sample. We additionally performed a sensitivity analysis for weeks of abstinence by including the weeks preceding the 9th rTMS-treatment visit and immediate post-visit since these two weeks represent the time following the delivery of a total of 16-sessions of rTMS (a similar dose to early rTMS for depression studies). We calculated the relative risk of having a week of abstinence as a measure of the effect size, along with 95% confidence intervals. We used creatinine-corrected urine cannabinoids to verify self-reported abstinence at follow-up weeks 2 and 4, and calculated the proportion that these two methods were concordant.

To evaluate group differences in the number of days of cannabis use (Aim-2b), we utilized a GLMM with the number of days-per-week of cannabis use as the dependent measure. We included participants in the model who had follow-up data (the completer sample; N=51) and given the apparent divergence in days-per-week of cannabis use in the final two-weeks of the follow-up period, we chose to focus our analysis on that period.

Residual normality was assessed for each of the above models using QQ-plots. We included sex in all models to detect a potential effect, but when no significant effects were found, we excluded sex from the subsequent final models. Apart from craving, this was a pilot trial and not powered a priori to detect statistically significant differences; however, statistically significant *p-*values are noted when observed. When reported, a level of α = 0.05 was used (two-tailed), and no adjustments for multiple comparisons were made for this preliminary investigation. Effect sizes at each time point were estimated as Cohen’s d using means and pooled standard deviations. Statistical analyses were conducted using R software (version 4.2.1, GNU project) and SAS version 9.4 (Cary, NC).

## 3.0 Results

### 3.1 Participants and Trial Feasibility (Figure-1 and Table-1)

**Figure-1:**
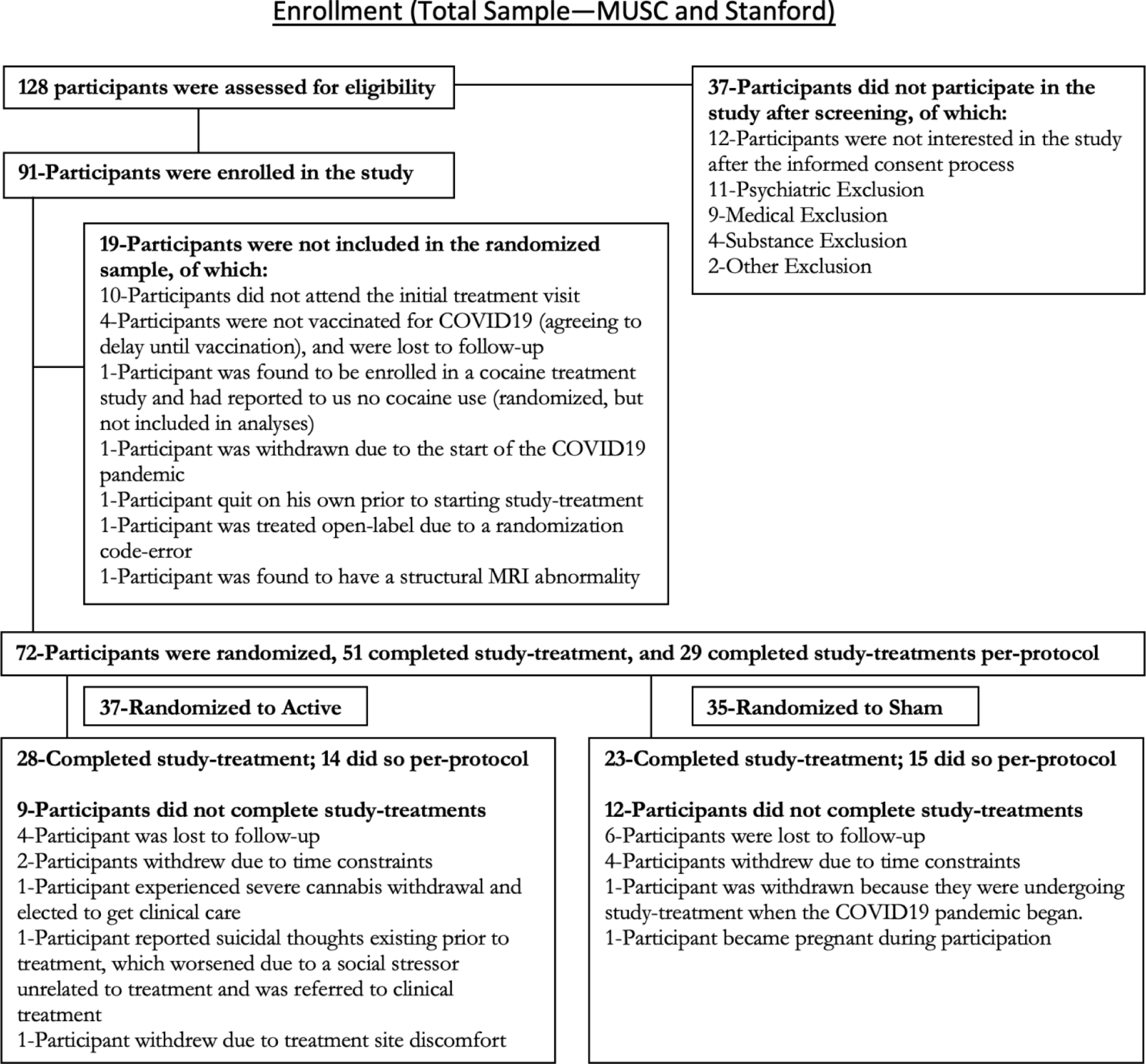
Consort Flow Diagram of the oaverall sample.

**Table-1:**
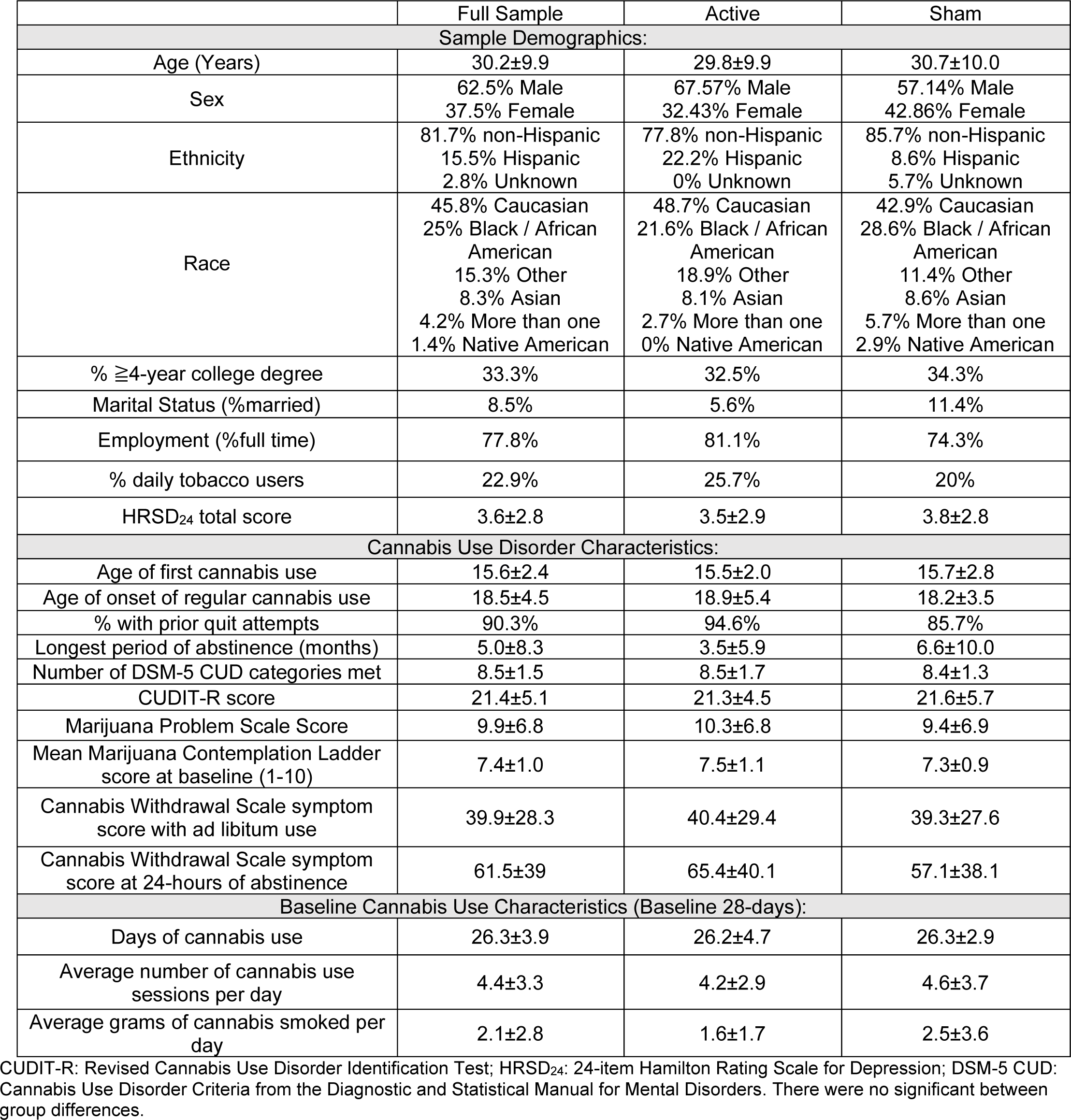
Baseline and demographic characteristics of the Intent to Treat (ITT) sample. All values are reported ± Standard Deviations. Cannabis use variables are reported for the 28-days prior to the screening and enrollment visit.

|  | Full Sample | Active | Sham |
| --- | --- | --- | --- |
| **Sample Demographics:** |  |  |  |
| Age (Years) | 30.2±9.9 | 29.8±9.9 | 30.7±10.0 |
| Sex | 62.5% Male<br>37.5% Female | 67.57% Male<br>32.43% Female | 57.14% Male<br>42.86% Female |
| Ethnicity | 81.7% non-Hispanic<br>15.5% Hispanic<br>2.8% Unknown | 77.8% non-Hispanic<br>22.2% Hispanic<br>0% Unknown | 85.7% non-Hispanic<br>8.6% Hispanic<br>5.7% Unknown |
| Race | 45.8% Caucasian<br>25% Black / African American<br>15.3% Other<br>8.3% Asian<br>4.2% More than one<br>1.4% Native American | 48.7% Caucasian<br>21.6% Black / African American<br>18.9% Other<br>8.1% Asian<br>2.7% More than one<br>0% Native American | 42.9% Caucasian<br>28.6% Black / African American<br>11.4% Other<br>8.6% Asian<br>5.7% More than one<br>2.9% Native American |
| % ≥4-year college degree | 33.3% | 32.5% | 34.3% |
| Marital Status (%married) | 8.5% | 5.6% | 11.4% |
| Employment (%full time) | 77.8% | 81.1% | 74.3% |
| % daily tobacco users | 22.9% | 25.7% | 20% |
| HRSD<sub>24</sub> total score | 3.6±2.8 | 3.5±2.9 | 3.8±2.8 |
| **Cannabis Use Disorder Characteristics:** |  |  |  |
| Age of first cannabis use | 15.6±2.4 | 15.5±2.0 | 15.7±2.8 |
| Age of onset of regular cannabis use | 18.5±4.5 | 18.9±5.4 | 18.2±3.5 |
| % with prior quit attempts | 90.3% | 94.6% | 85.7% |
| Longest period of abstinence (months) | 5.0±8.3 | 3.5±5.9 | 6.6±10.0 |
| Number of DSM-5 CUD categories met | 8.5±1.5 | 8.5±1.7 | 8.4±1.3 |
| CUDIT-R score | 21.4±5.1 | 21.3±4.5 | 21.6±5.7 |
| Marijuana Problem Scale Score | 9.9±6.8 | 10.3±6.8 | 9.4±6.9 |
| Mean Marijuana Contemplation Ladder score at baseline (1-10) | 7.4±1.0 | 7.5±1.1 | 7.3±0.9 |
| Cannabis Withdrawal Scale symptom score with ad libitum use | 39.9±28.3 | 40.4±29.4 | 39.3±27.6 |
| Cannabis Withdrawal Scale symptom score at 24-hours of abstinence | 61.5±39 | 65.4±40.1 | 57.1±38.1 |
| **Baseline Cannabis Use Characteristics (Baseline 28-days):** |  |  |  |
| Days of cannabis use | 26.3±3.9 | 26.2±4.7 | 26.3±2.9 |
| Average number of cannabis use sessions per day | 4.4±3.3 | 4.2±2.9 | 4.6±3.7 |
| Average grams of cannabis smoked per day | 2.1±2.8 | 1.6±1.7 | 2.5±3.6 |

CUDIT-R: Revised Cannabis Use Disorder Identification Test; HRSD<sub>24</sub>: 24-item Hamilton Rating Scale for Depression; DSM-5 CUD: Cannabis Use Disorder Criteria from the Diagnostic and Statistical Manual for Mental Disorders. There were no significant between group differences.

We assessed a total of 128 participants for eligibility between the two sites, enrolled 91 during the screening visit, and randomized 72 during an initial study treatment visit (comprising the Intent-To-Treat—ITT—sample). Fifty-one participants completed all study visits (comprising the completer sample). Participants tolerated study-treatment well (mean final treatment dose 114.7%±9.4%rMT-active; 118.9%±3.7%rMT-sham). There were no statistically significant differences between conditions on any baseline variable. There were, however, several site differences that reached statistical significance (Supplemental Table-1), including race and ethnicity, number of daily tobacco users, total HRSD_24_ score, number of DSM-5 criteria for CUD, MPS total score, number of grams of cannabis smoked per day, and number of days using cannabis in the baseline 28-days.

### 3.2 Cannabis Craving (Figure-2)

**Figure-2:**
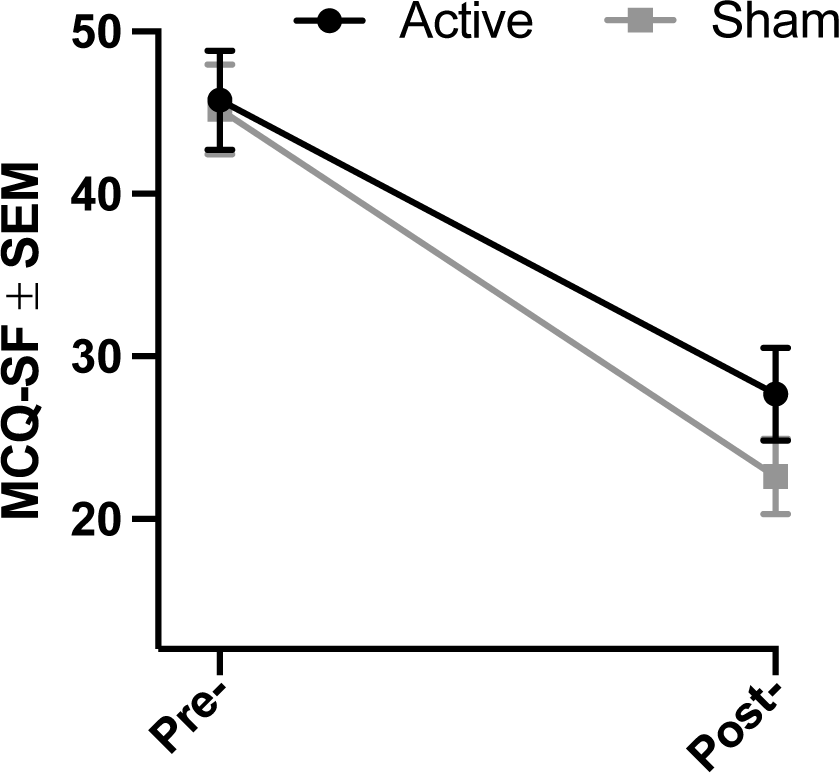
Change in Craving Pre- and Post- Treatment Course. Craving as measured by the short form of the Marijuana Carving Questionnaire (MCQ-SF), before and after the delivery of the full course of rTMS. Mean MCQ-SF scores are reported with Standard Errors of the Means (±SEM).

Craving as measured by the MCQ-SF total score decreased in both active and sham conditions between pre- and post-treatment course assessments (45.8±18.5SD to 27.7±15.1SD in the active-group; 45.2±16.3SD to 22.6±11.1SD in the sham-group; active vs. sham: 4.05; Z=0.98, *p*=0.33 when adjusting for site). None of the exploratory covariates were significantly related to the change in the MCQ-SF score and minimally changed the treatment effects.

### 3.3 Abstinence from Cannabis (Figure-3 and Supplemental Table-2)

**Figure-3:**
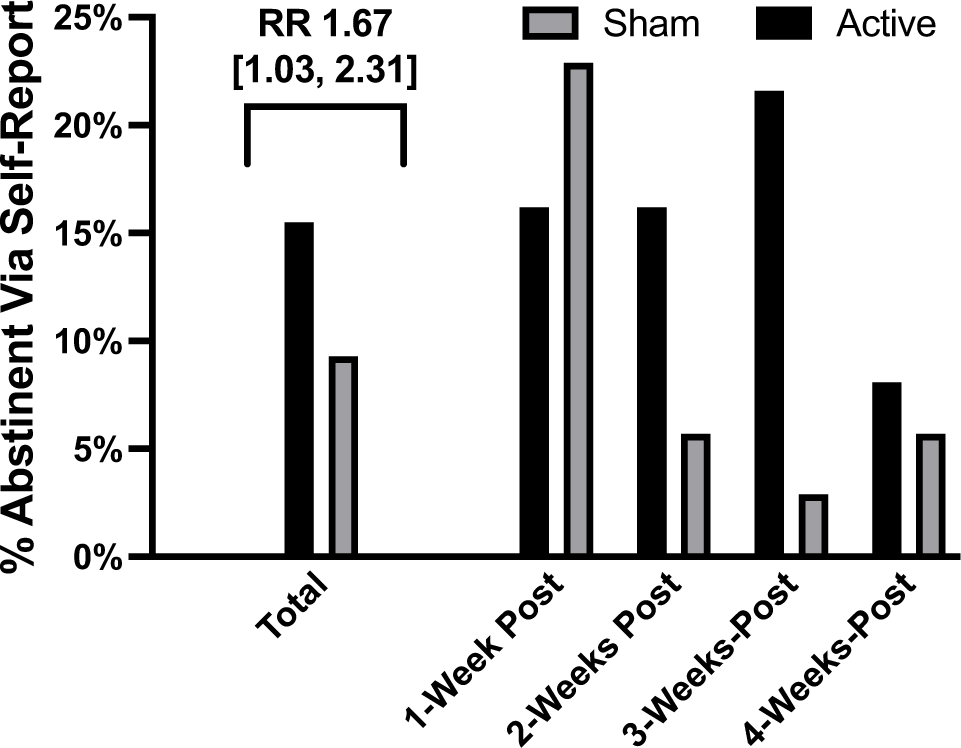
Weeks of Self-Reported Abstinence. This figure depicts the number of self-reported weeks of abstinence. For this intent to treat sample missing data is imputed as non-abstinent. The ‘total’ percent weeks of abstinence is reported as the Relative Risk of having a week of abstinence in the follow-up period with the 95% confidence interval. Of note, when modeling between group differences in the four weeks of follow-up, and adjusting for site, the rate ratio of 1.66 does not meet statistical significance [95% CI: 0.84, 3.28]; *p*=0.14.

Participants who received active stimulation reported a numerically higher percent of weeks of abstinence in the four-week follow-up period (15.5%) than those who received sham stimulation (9.3%) with a relative risk of having a week of abstinence of 1.67 [95%CI: 1.03, 2.31]. However, the average number of weeks of abstinence in the 4-week follow-up period did not significantly differ between the two-conditions when adjusting for the site (rate ratio = 1.66 [95% CI: 0.84, 3.28]; *p*=0.14). Of note, much of the abstinence effect was driven by the Stanford sample (31.3%-active; 15.0%-sham; rate ratio=2.08 [95% CI: 0.95, 4.58]; p=0.07), with minimum abstinence in either the active or sham group in the MUSC sample (3.6%-active; 5.0%-sham; rate ratio=0.71 [95% CI: 0.16, 3.19]; p=0.66). Creatinine-corrected urine cannabinoids were concordant with self-reported abstinence in 84.6% of observations. The number of weeks of self-reported abstinence was similar in the final 6-weeks of the study (14.9%-active, 9.0%-sham; rate ratio for active vs. sham = 1.63 [95% CI: 0.93, 2.87]; *p*=0.09).

### 3.4 Days-Per-Week of Cannabis Use (Figure-4)

**Figure-4:**
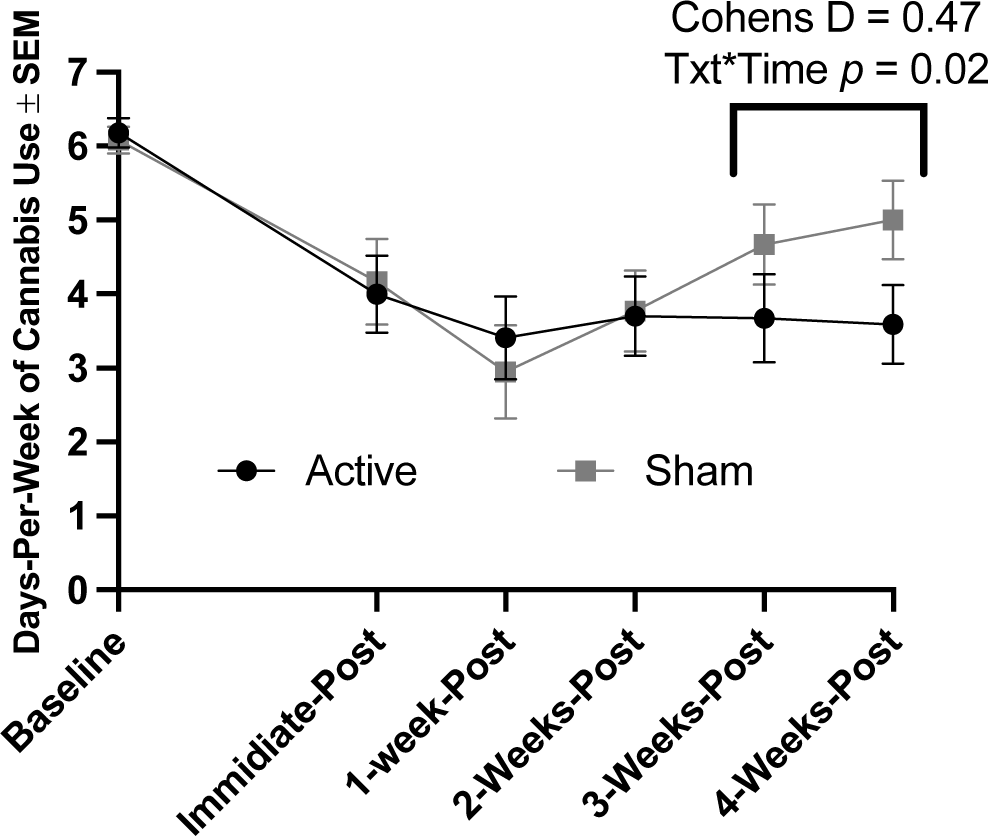
Days-Per-Week of Cannabis Use. Days per week of cannabis use: This chart represents the number of days any cannabis was used in the preceding week. Scores are reported with Standard Errors of the Means (±SEM).

The mean number of days-per-week of cannabis use decreased in both conditions between the pre-intervention week and the four weeks of follow-up but diverged between the active and sham groups in the final two-weeks of follow-up. The active group appeared to have a more durable decrease in days-per-week of cannabis use (6.0±1.2SD-pre, 4.0±2.7SD-post, 3.4±2.9SD-1-week-FU, 3.7±2.8SD-2-week-FU, 3.7±3.1SD-3-week-FU, and 3.6±2.8SD-4-week-FU) than did the sham group (6.0±1.4SD-pre, 4.0±2.8SD-post, 2.8±3.0SD-1-week-FU, 3.8±2.5SD-2-week-FU, 4.8±2.5SD-3-week-FU, and 5.1±2.4SD-4-week-FU). The reduction in days-per-week of cannabis use from follow-up week two to follow-up week four differed significantly by treatment condition (active vs. sham: -0.72; Z=-2.33, *p*=0.02). The effect sizes, as measured by Cohen’s d during the post-treatment divergence, were 0.39 in week 3, 0.57 in week 4 and 0.47 when combining the final two weeks. The Stanford sample again had a larger effect on days-per-week of use in the final week of the follow-up period (Cohen’s d=0.72), though a small effect was also present in the MUSC sample (Cohen’s d=0.34).

### 3.5 Adverse Events (AEs—Table-2)

We included all adverse events (AEs) that were rated as definitely, probably, possibly, and probably not related to the study intervention in the analysis. A total of 30 AEs occurred in 23 participants. There were no severe or serious AEs. Headache was the most common AE, followed by fatigue, and a series of AEs that occurred only once. Twenty-nine of the 30 AEs were categorized as mild and one as moderate (a participant in the sham group experienced a more substantial headache). Two participants (both in the active condition) described a clear onset of symptoms consistent with the cannabis abstinence syndrome^63^ coinciding temporally with when they stopped using cannabis. We coded these clusters of AEs as definitely not related to study-treatment and did not include them in the AE analysis. Other AEs that were more likely to have been withdrawal related than rTMS related (including insomnia, irritability/mood swings, and increased anxiety) were included in the AE analysis, given they were singular symptoms (as opposed to clearly clustered symptoms).

### 3.6 Blinding (Supplemental Table-3)

Following the first rTMS study visit, 91.2% of the active and 73.5% of the sham groups believed they received active rTMS (*p*=0.11). Following the full course of rTMS, 61.5% of the active and 60.9% of the sham groups believed they received active rTMS (*p*=0.96).

## 4.0 Discussion

In this two-site, phase-2, randomized, sham-controlled clinical trial, we demonstrated the feasibility of delivering an rTMS intervention along with three-sessions of motivational enhancement therapy to treatment-seeking participants with moderate or severe CUD. Participants tolerated rTMS well with few adverse events, and the retention rate in this study compares well to other CUD studies. Contrary to our hypothesis, we did not find between group differences in craving reduction. However, both groups experienced a reduction in craving scores. We observed numerically (but not statistically significantly) more weeks of abstinence in the active rTMS group than the sham rTMS group in the follow-up period, driven mainly by a large effect in the Stanford sample. The number of days-per-week of cannabis use decreased during active treatment in both groups, though remained reduced in the follow-up period in the active group but not in the sham group, an effect that differed significantly in the final two-weeks of follow-up.

As expected, rTMS was well tolerated with only mild AEs observed in the active condition. The AE profile of rTMS compares favorably to those described in studies using medications reporting a positive effect, including quetiapine, varenicline, and, nabiximols^11–13^. Similarly, the overall retention rate in this trial compared favorably to other clinical trials, with 70.8% of the sample retained (retention rates for pharmacotherapy trials have ranged from 36% to 65%^11–13,64–70^). Notably, the active group had a higher retention rate than the sham group suggesting that study treatment was well tolerated. Blinding was intact for this study and we believe the sham procedures can be relied upon for future investigations.

Most but not all studies across substance use disorders have suggested that rTMS applied to the left-DLPFC reduces craving^71^. As such, it is surprising that the participants within the sham group of our trial reported numerically decreased craving relative to the active group. Our initial explanation for this unexpected finding was that a covariate must have driven the post-treatment craving score. However, in exploratory modeling, none of the final models significantly predicted the final MCQ-SF score, and the inclusion of candidate covariates minimally changed the models. The limited power available in our medium-sized sample, however, may have reduced the ability of the explored covariates from becoming significant predictors during modelling. Another possibility is that the focus and timeframe (the immediate time of its administration) of the MCQ-SF limited its ability to detect between-group differences in the context of this clinical trial. Notably, no pharmacotherapy trial (including full agonist therapy^11,65^) has observed a between-group change in craving over the course of treatment using the MCQ-SF, even when finding a clinical effect. The MCQ-SF subsequently may be better at detecting acute differences^38,57^ than longitudinal ones. Future clinical trials might consider including a measure that captures a longer time period and has shown sensitivity to treatment effects in other substance use disorders.

The moderate between-group clinical effect-sizes we observed in the follow-up period of our trial were in the range of, or larger, than other treatment trials observing suggestions of efficacy in CUD. The between-group relative risk of having a week of abstinence in the present trial approached but did not meet the observed effect in our recent phase-2 varenicline trial^13^. Self-reported weeks-of-abstinence were verified using cannabinoid testing in nearly 85% of cases, supporting the validity of participants reporting abstinence. The between-group effect size of days-per-week of cannabis use of Cohen’s d=0.47 was comparable to the two pharmacotherapy trials that observed a difference in days-per-week of use (Cohen’s d of 0.39-0.55)^11,13^, and to behavioral trials using a wait-list group as a comparator (Hedges’ g of 0.44 in meta-analysis)^14^. Of note, though we did hypothesize that participants receiving active rTMS would have fewer days of cannabis use in the follow-up period, our observed effect did not emerge until the final two-weeks of follow-up, which was not pre-hypothesized. All participants received an evidence-based three-session course of MET and had a high rate of clinical contact with supportive clinical staff, which in addition to placebo effects may explain the similar clinical improvement in both groups during the acute treatment period. The divergence in days-per-week of cannabis use subsequently occurred when the intensity of clinical visits was reduced and could represent an increased durability in the effect following treatment. Two other recent sham-controlled rTMS for alcohol use disorder trials observed a similar phenomenon where the main treatment-effects were reflected in increased durability in the follow-up period^34,35^. Future rTMS for addiction trials might focus further on durability in an extended follow-up period.

Our study sample differed by site in several baseline variables and the preliminary effect size of the therapeutic effects of rTMS also differed by site, most notably in the number of weeks-of-abstinence. Both samples heavily used cannabis at baseline; however, the sample in California reported more days-per-week of use and numerically more use sessions per day at baseline. The group in South Carolina reported smoking more grams-per-day of cannabis and more co-use of cigarettes. This differential use pattern may be more consistent with the use of high-potency cannabis in the California sample, which would paradoxically result in using fewer grams of cannabis (i.e., higher-potency cannabis would require a lower dose to get a similar or larger effect than low-potency cannabis). Indeed, such a use pattern has been previously described as a differentiator of cannabis use in states where marijuana is legal vs. illegal — cannabis users in ‘legal’ states are more likely to employ higher potency methods of cannabis use (dabs, vapes, etc.)^72,73^ than in ‘illegal’ states. Higher potency cannabis and vape/dab use means are associated with more cannabis-related problems^74–76^ and would be consistent with the higher average marijuana problem scale score and more DSM-5 CUD criteria met in the California sample. These site differences may account for the differential treatment effects as more impaired participants reporting more marijuana-related problems may have been more motivated to reduce the amount of cannabis they use. Non-specific site effects are also possible, though unlikely given that the study’s principal investigator (GLS) was responsible for the conduct of the trial at both study sites. Time effects are also possible, especially since the COVID19 pandemic began just as the study transitioned from South Carolina to California (the sample that was recruited in California was enrolled after the onset of the pandemic, whereas the sample recruited in South Carolina was enrolled before the pandemic). Cannabis use increased broadly during the pandemic, and the isolation inherent in the pandemic potentially increased problematic cannabis use, making pandemic effects a possible contributor to the different populations^77^. Regardless of the site effects, there were suggestions of treatment efficacy at both sites, particularly in the important variable days-per-week of cannabis use. It is possible that the California participants with more severe CUD were motivated more for abstinence, and the South Carolina participants with less severe CUD were satisfied with reducing the number of days they used cannabis.

Several limitations of this study merit mentioning to add context to our findings. Methodologically, during data collection, we did not differentiate various means of cannabis use, and so in our data analysis, we were not able to differentiate participants with different methods of cannabis use. Given our initial concerns about retention rates^39^, we set the number of rTMS sessions (twenty total delivered over 10-visits), the density of visits (two-per-week), and inter-session interval (30-minutes), at the minimums we thought would have the potential to have an effect. We also used scalp-based targeting (Beam-F3) as opposed to more sophisticated targeting, such as MRI guided targeting, for feasibility and dissemination purposes. All of these compromises, though successful in terms of trial feasibility, may have resulted in a lower efficacy rate relative to a more optimized treatment paradigm including additional study-treatments^78^, an increased inter-session interval^79^, and a higher density of visits per week. Further, as this was a sequential rather than concurrent multi-site study, it is unclear if treatment by site interactions were a result of site differences or time differences. Finally, our analysis of days-per-week of cannabis use in the final two-weeks of follow-up was not pre-planned or corrected for multiple comparisons so we would categorize that finding as more hypothesis-generating for future work than hypothesis-proving.

## 5.0 Conclusions and Future Directions

In summary, these preliminary findings suggest rTMS applied to the DLPFC holds promise in assisting participants with CUD to reduce their cannabis use, though further study with an optimized rTMS treatment paradigm and a longer follow-up period is needed to determine whether, in fact, such a treatment paradigm can develop into a standard of care intervention. Further study is warranted given the excellent safety profile found in this study, the high retention rate (demonstrating feasibility), and the promising clinical effect-sizes observed despite the relatively low dose of rTMS administered.

**Table 2:** Adverse Events between groups.

|  | Total Sample, N,<br>and (% of ITT<br>sample—total / 72) | Active condition,<br>N, and (% of ITT<br>sample—total / 37) | Sham condition,<br>N, and (% of ITT<br>sample—total / 35) | Significance |
| --- | --- | --- | --- | --- |
| # of participants with Any<br>Adverse Events (%) | 23 (31.9%) | 16 (43.2%) | 7 (20.0%) | $p=0.04$ |
| Total number of Adverse<br>events | 30 | 19 | 11 | $p=0.05$ |
| Headache (%) | 14 (19.4%) | 10 (27.0%) | 4 (11.4%) | *ns* |
| Fatigue (%) | 9 (12.5%) | 6 (16.2%) | 3 (8.6%) | *ns* |
| Eye Twitch (%) | 1 (1.4%) | 1 (2.7%) | 0 (0) | *ns* |
| Jaw Pain (%) | 1 (1.4%) | 1 (2.7%) | 0 (0) | *ns* |
| Site discomfort (%) | 1 (1.4%) | 1 (2.7%) | 0 (0) | *ns* |
| Insomnia (%) | 1 (1.4%) | 0 (0) | 1 (2.9%) | *ns* |
| Irritability / mood swings (%) | 1 (1.4%) | 0 (0) | 1 (2.9%) | *ns* |
| Hand numbness (%) | 1 (1.4%) | 0 (0) | 1 (2.9%) | *ns* |
| Increased anxiety (%) | 1 (1.4%) | 0 (0) | 1 (2.9%) | *ns* |

## Data Availability

All data produced in the present study are available upon reasonable request to the authors.

## Author Contributions

GLS, NLB, EBS, TKK, MSG, and ALM designed the experiment. GLS, BLW, MAC, LAC, IK, BJS, TJF, AHM, NRW, AJM, IHK, EBS, and TKK performed the study. GLS, BK, NLB, and JPK analyzed the study’s findings. GLS, BK, NLB, JPK, and ALM wrote the first draft of the manuscript. All authors critically reviewed and edited the manuscript.

**Clinicaltrials.gov identifiers:** NCT03144232.

## Acknowledgments

We would like to acknowledge the many contributors who made this study possible, including Amanda Wagner, Taylor Rodgers, Robert Malcolm, Lisa Nunn, Nick Bassano, and Alan Schatzberg.

## Supplemental Tables and Figures

**Supplemental Figure-1:**
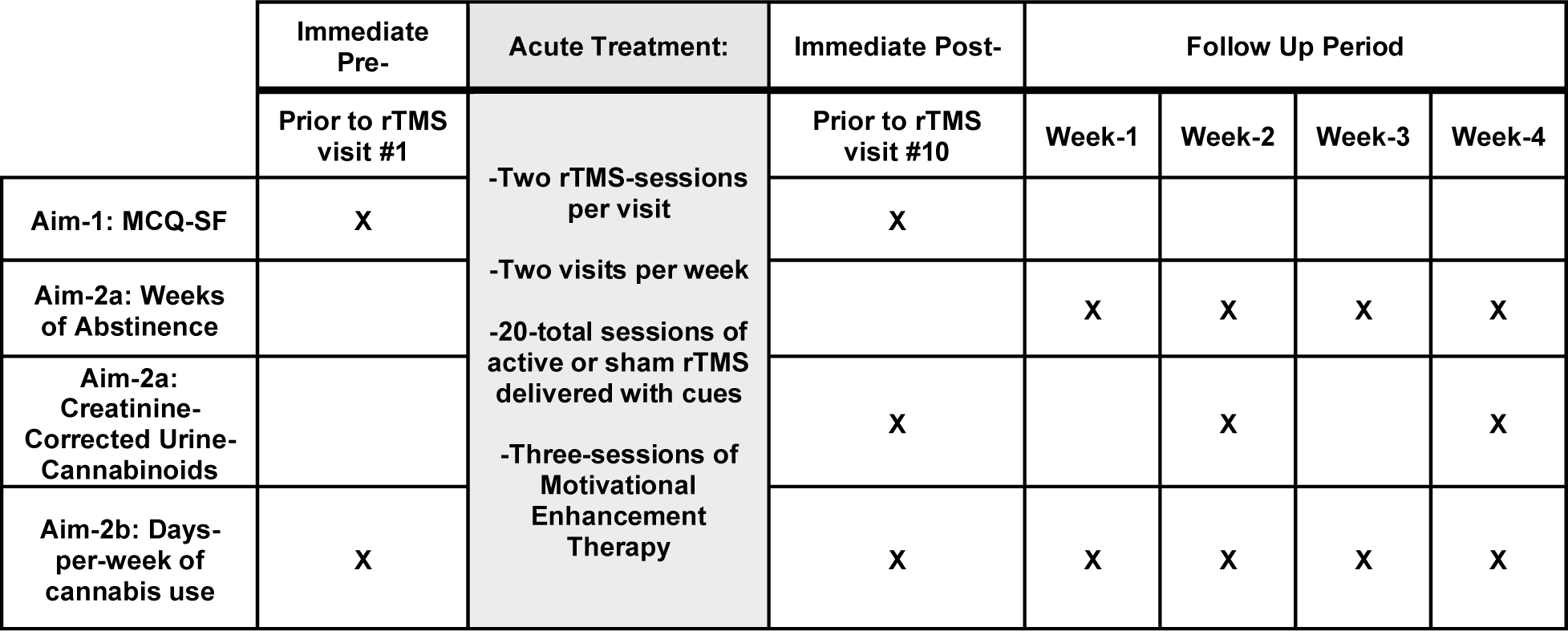
Experimental Timeline of Treatments and Assessments:

**Supplemental Table-1:** Demographics comparison between Stanford and MUSC samples. Baseline and demographic characteristics in the Intent to Treat (ITT) sample between the MUSC and Stanford sites. All values are reported ± Standard Deviations. Cannabis use variables are reported for the 28-days prior to the screening and enrollment visit.

|  | Full Sample | MUSC | Stanford | Significance |
| --- | --- | --- | --- | --- |
| **Sample Demographics:** |  |  |  |  |
| Age (Years) | 30.2±9.93 | 31.4±10.3 | 28.7±9.4 | ns |
| Sex | 62.5% Male<br>37.5% Female | 34.1% Female<br>65.9% Male | 41.9% Female<br>58.1% Male | ns |
| Ethnicity | 81.7% non-Hispanic<br>15.5% Hispanic<br>2.8% Unknown | 92.5% non-Hispanic<br>2.5% Hispanic<br>5% Unknown | 67.7% non-Hispanic<br>32.3% Hispanic<br>0% Unknown | $p=0.0008$ |
| Race | 45.8% Caucasian<br>25% Black / African American<br>15.3% Other<br>8.3% Asian<br>4.2% More than one<br>1.4% Native American | 56.1% Caucasian<br>36.6% Black / African American<br>2.4% Other<br>0% Asian<br>2.4% More than one<br>2.4% Native American | 33.3% Caucasian<br>9.7% Black / African American<br>32.3% Other<br>19.4% Asian<br>32.3% more than one<br>0% Native American | $p=0.0001$ |
| % ≥4-year college degree | 33.4% | 31.7% | 35.5% | ns |
| Marital Status (%married) | 8.5% | 7.5% | 9.7% | ns |
| Employment (%full time) | 77.8% | 78.1% | 77.4% | ns |
| % daily tobacco users | 22.9% | 35.9% | 6.5% | $p=0.004$ |
| **Cannabis Use Disorder Characteristics:** |  |  |  |  |
| Age of first cannabis use | 15.6±2.4 | 15.3±2.5 | 16.0±2.3 | ns |
| Age of onset of regular cannabis use | 18.5±4.5 | 19.0±5.6 | 17.9±2.5 | ns |
| % with prior quit attempts | 90.3% | 92.7% | 87.1% | ns |
| Longest period of abstinence (months) | 5.0±8.3 | 4.6±9.1 | 5.6±7.2 | ns |
| Number of DSM-5 CUD categories met | 8.5±1.5 | 7.9±1.5 | 9.2±1.2 | $p=0.001$ |
| CUDIT-R score | 21.4±5.1 | 20.9±4.8 | 22.2±5.4 | ns |
| Marijuana Problem Scale Score | 9.9±6.8 | 7.3±5.6 | 13.2±6.8 | $p=0.0001$ |
| Mean Marijuana Contemplation Ladder score at baseline (1-10) | 7.4±1.0 | 7.5±1.1 | 7.3±1.0 | ns |
| Cannabis Withdrawal Scale symptom score with ad libitum use | 39.9±28.3 | 34.3±27.1 | 42.9±28.9 | ns |
| Cannabis Withdrawal Scale symptom score at 24-hours of abstinence | 61.5±39.0 | 56.2±41.3 | 64.6±37.9 | ns |
| Baseline Cannabis Use Characteristics (Baseline 28-days): |  |  |  |  |
| Days of cannabis use (last 28-days) | 26.3±3.9 | 25.4±4.8 | 27.4±1.7 | $p=0.01$ |
| Average number of cannabis use sessions/day | 4.4±3.3 | 3.8±3.2 | 5.2±3.4 | ns |
| Average grams cannabis smoked/day | 2.1±2.8 | 2.7±3.4 | 1.2±1.5 | $p=0.02$ |

CUDIT-R: Revised Cannabis Use Disorder Identification Test; HRSD<sub>24</sub>: 24-item Hamilton Rating Scale for Depression; DSM-5 CUD: Cannabis Use Disorder Criteria from the Diagnostic and Statistical Manual for Mental Disorders

**Supplemental Table-2:** Integrity of the Blind

| Group: | Guessed Active after first visit: | Confidence (0-Not confident at all, 4-Almost Certain) | Guessed Active after last visit: | Confidence: (0-Not confident at all, 4-Almost Certain) | Significance |
| --- | --- | --- | --- | --- | --- |
| Active Group | 91.2% | 1.3±1.2 | 61.5% | 2.0±1.3 | $p=0.11$ First |
| Sham Group | 73.5% | 1.6±1.1 | 60.9% | 1.7±1.5 | $p=0.96$ Last |
| Active MUSC | 94.4% | 1.3±1.2 | 45.5% | 2.0±1.3 | $p=0.09$ First |
| Sham MUSC | 68.4% | 1.6±1.1 | 60.0% | 1.7±1.5 | $p=0.67$ Last |
| Active Stanford | 87.5% | 1.3±1.0 | 73.3% | 1.9±1.3 | $p=0.65$ First |
| Sham Stanford | 80.0% | 1.5±1.2 | 61.5% | 1.7±1.6 | $p=0.69$ Last |

**Supplemental Table-3:** Weeks of abstinence: Percent of participants in the intent to treat (ITT) sample who reported no cannabis use sessions over a week.

| Full sample: | Self-Report abstinence (ITT) |  | Abstinence as verified by Urine Drug Testing (ITT) |  | Self-Report abstinence (Available Data) |  | Abstinence as verified by Urine Drug Testing (Available) |  |
| --- | --- | --- | --- | --- | --- | --- | --- | --- |
| Condition: | Active | Sham | Active | Sham | Active | Sham | Active | Sham |
| Total Tx9-4wk-FU: | 14.9%<br>(33/222) | 9.0%<br>(19/210) | 10.8%<br>(12/111) | 5.7%<br>(6/105) | 20.4%<br>(33/162) | 13.9%<br>(19/137) | 14.8%<br>(12/81) | 8.8%<br>(6/68) |
| Total F/U period only | 15.5%<br>(23/148) | 9.3%<br>(13/140) | 10.8%<br>(8/74) | 4.3%<br>(3/70) | 21.3%<br>(23/108) | 14.4%<br>(13/90) | 14.8%<br>(8/54) | 6.8%<br>(3/44) |
| Treatment visit 9 | 13.5%<br>(5/37) | 5.7%<br>(2/35) |  |  | 18.5%<br>(5/27) | 8.7%<br>(2/23) |  |  |
| Immediate post | 13.5%<br>(5/37) | 11.4%<br>(4/35) | 10.8%<br>(4/37) | 8.6%<br>(3/35) | 18.5%<br>(5/27) | 16.7%<br>(4/24) | 14.8%<br>(4/27) | 12.5%<br>(3/24) |
| 1-week FU | 16.2%<br>(6/37) | 22.9%<br>(8/35) |  |  | 22.2%<br>(6/27) | 34.8%<br>(8/23) |  |  |
| 2-week FU | 16.2%<br>(6/37) | 5.7%<br>(2/35) | 13.5%<sup>#</sup><br>(5/37) | 2.9%<sup>#</sup><br>(1/35) | 22.2%<br>(6/27) | 8.7%<br>(2/23) | 18.5%<br>(5/27) | 4.5%<br>(1/22) |
| 3-week FU | 21.6%*<br>(8/37) | 2.9%*<br>(1/35) |  |  | 29.6%<sup>^</sup><br>(8/27) | 4.6%<sup>^</sup><br>(1/22) |  |  |
| 4-week FU | 8.1%<br>(3/37) | 5.7%<br>(2/35) | 8.1%<br>(3/37) | 5.7%<br>(2/35) | 11.1%<br>(3/27) | 9.1%<br>(2/22) | 11.1%<br>(3/27) | 9.1%<br>(2/22) |

